# Self-medication with antibiotics- a survey among traders in the central business district of Accra, Ghana

**DOI:** 10.1101/2021.10.31.21265726

**Authors:** S. K. Ofori, E.A. Akowuah, C.E. Amankwa, D. Babatunde, F. Baiden

## Abstract

**Background:** The misuse of antibiotics is a global problem, and the form of misuse varies among different countries and cultures. The study explored antibiotic self-medication practices among traders in Accra, Ghana.

**Methods:** A paper-based questionnaire survey was conducted among traders found in a convenient sample of shops in the Central Business District (CBD) between November 2016 and January 2017. The data was analyzed with Stata version 14.0. Antibiotic self-medication was defined as the use of antibiotics without the prescription of a recognized practitioner within 12 months of the survey. Descriptive analysis and analytical statistics using multivariate logistic regression analysis were performed to identify the factors associated with antibiotic self-medication.

**Results:** Four hundred and seventeen (417) traders (60.0% females) with a mean age of 36 (±10.6) years were surveyed. The prevalence of antibiotic self-medication was 66.7%. Penicillins were the most misused. Upper respiratory infections and gastrointestinal tract infections were the leading reasons for antibiotic self-medication. The most common sources of information on self-medicated antibiotics were community pharmacies (55.2%), doctors (17.8%), and package inserts (17.2%). People with tertiary education were less likely to use antibiotics for self-medication compared to those with no education (OR=0.39 (95% C.I; 0.16-0.98). Persons who sold food items (aOR = 3.56, 95% CI; 1.70-7.45), cosmetics (aOR = 2.61, 95% CI; 1.34-5.09), and clothing (aOR= 3.39, 95% CI; 1.88-6.12) were more likely to use antibiotics for self medication compared to those who sold other items.

**Conclusion:** Self-medication with antibiotics was high among traders in the CBD of Accra, Ghana. Traders need to be educated on the proper home management of URTIs. Measures are required to restrict their access to antibiotics for self-medication.

## Background

Self-medication with antibiotics is a global public health problem with major implications for developing countries [1, 2]. An increase in drug resistance has led to an exponential rise in th cost of treatment for infections, treatment failures, and increased mortality from life-threatenin infectious diseases [3]. One of the driving determinants of self-medication with antibiotics is the ease of access over the counter due to poor regulations, and this leads to the use of antibiotics for wrong indications, drug interactions, and adverse effects from contraindications [4]. Furthermore, evidence suggests that people in developing countries receive their drugs as well as information on these drugs from non-pharmacists and non-trained personnel [5]. Information may also be obtained from informal sources, including friends, family [6], and the media [7].

Overuse and inappropriate prescribing of broad-spectrum antibiotics is a driving factor of antibiotic resistance. Studies have reported that at least half of prescribed antibiotics are not necessary or are incorrectly chosen. [8, 9] This leads to an increase in leftover drugs at homes which serves as easy access when one experiences similar symptoms. Watkins and colleagues assessed the community perceptions of antibiotic access and use and reported that participants who stopped taking antibiotics because they felt better kept leftovers [10].

A study in Egypt reported the prevalence rate of antibiotic abuse as 53.9% and mentioned cough and common cold as the symptoms that warranted the use of antibiotics [7]. Another study in Ethiopia reported a prevalence of 8.4%, which was far lower than that recorded in Egypt [11]. In Nigeria, a prevalence of 53.8 % was reported among university students who admitted to taking antibiotics for self-medication [12].

As found in South Africa, self-medication is a means by which people avoid paying hospital fees and overcome other barriers to appropriate health care [5, 10]. Self-medication with drugs that include antibiotics has been reported in Ghana. Although a policy exists for regulating all medicines in Ghana, the control of antibiotics use is not covered in the National Drug Policy, which makes it easier for antibiotics to be accessed by the community. Persons eligible to prescribe antibiotics in Ghana include medical doctors, physician assistants, midwives, an trained nurses depending on the type of antibiotics [13, 14]. However, the prescription and dispensing by ineligible individuals promote the inappropriate use of antibiotics for self-medication, especially among vulnerable groups [13].

Previous studies on self-medication practices in Ghana have focused on groups in the country’s formal sector. This includes tertiary students, medical students, and health practitioner[12, 15-17]. Ghana has a large informal sector that is dominated by petty trade and subsistence. Interventions to promote the appropriate use of antibiotics in Ghanaian society will require a good understanding of practices among people in the country’s informal sector. Therefore, this study aimed to explore antibiotic self-medication practices among traders, including estimating the prevalence and identifying associated factors.

## Method

### Study Population

The study was conducted in the Central Business District (CBD) of Accra. The markets are dominated by women who sell fresh food, imported goods, local jewelry, shoes, and many more. The site was chosen because it is one of the busiest places in Accra, and it has a popular area within it called “Drug Lane” where many retail pharmacy shops are located. Participants were selected using convenient sampling. This is because traders are mobile, and it was not possible to obtain a good sampling frame. One person was chosen from each shop/stall to avoid the cluster effect.

### Data collection techniques and tools

A paper-based structured questionnaire was used for data collection. Data were collected between November 2016 and January 2017. Questions were close-ended with options for participants to choose from, and participants completed the questionnaires with the aid of trained research assistants. The questionnaire consisted of socio-demographic characteristics of study participants, types of illnesses for which antibiotics were sought, reasons for self-medication, sources of advice, and classes of antibiotics used for self-medication. Self-medication with antibiotics was defined as the use of antibiotics without prescription by a recognized practitioner within one year of the survey. Research assistants were trained to show samples of antibiotics to respondents.

### Sample size calculation

The study aimed to interview 417 traders under the assumption that it will afford an estimation of the prevalence of antibiotic self-medication within a margin of error of 4.3% at a 95% confidence level, using an assumed self-medication prevalence of 27.6% (18)

### Statistical Analysis

The data were entered using Microsoft Excel 2013 and analyzed using STATA, version 14.0. Descriptive statistical analysis was carried out to obtain summary tables and graphs containing the demographic characteristics of the study participants. A multivariate logistic regression analysis was used to adjust for the effects of all predictors shown to be significantly associated with the misuse of antibiotics for self-medication at the univariate logistic regression level. Odds ratios (ORs) were reported with their 95% confidence intervals (C. Is), with the level of statistical significance set at p<0.05 for all tests. Results were expressed as means and standard deviations for continuous variables. Categorical data were presented as frequencies and percentages, and a Chi-square test was used to test the differences. All confounding variables were catered for, and the effect of prominent associations was evaluated using a multivariate logistic regression model to obtain the adjusted odds ratios.

## Results

Four hundred and seventeen traders were surveyed between November 2016 and January 2017. Most traders (N=253, 60.7%) were females, and the average age was 35.58 years (±10.60). About half of them were married (N= 197 (47.2%)) and had secondary education (N= 222 (53.2%)).

### Types of antibiotics and indications for use among traders in CBD

The majority (N= 279 (66.7%)) of traders admitted antibiotic self-medication (Table 2). About a quarter (23.1%) had self-medicated once in the preceding 12 months, while 30.1% had done so twice. The primary indication for antibiotic self-medication were upper respiratory tract infections (33.7%)) such as sore throat, cold, catarrh, and cough, and gastrointestinal disorders like diarrhea and stomach pain (N=86 (30.8%)).

**Table 1:** Demographic characteristics of traders interviewed (N=417)

| Characteristics |  | Number of persons (%) |
| --- | --- | --- |
| Sex | Male | 164 (39.3) |
|  | Female | 253 (60.7) |
| Age (years) | Average | 35.58 ± 10.60(17-72) |
| Age (group) | 17- 30 | 161 (38.6) |
|  | 31-45 | 187 (44.8) |
|  | >45 | 69 (16.6) |
| Marital status | Single | 187 (44.8) |
|  | Married/ cohabiting | 197 (47.2) |
|  | Separated/ divorced | 15 (3.6) |
|  | widowed | 18 (4.4) |
| Ethnicity | Akan | 185 (44.4) |
|  | Ga | 129 (30.9) |
|  | Ewe | 71 (17.0) |
|  | Non-Ghanaian | 14 (3.4) |
|  | Other | 18 (4.3) |
| Educational level | None | 43 (10.3) |
|  | Primary | 96 (23.0) |
|  | Secondary | 222 (53.2) |
|  | Tertiary | 56 (13.4) |
| Religion | None | 4 (1.0) |
|  | Christian | 377 (90.4) |
|  | Muslim | 33 (8.6) |
| Items traded in | Food | 71 (17.0) |
|  | Cosmetics | 78 (18.7) |
|  | Clothing | 142 (34.1) |
|  | Stationary | 32 (7.7) |
|  | Other* | 94 (22.5) |
| Place of trade | Own shop | 149 (35.7) |
|  | Rented shop | 128 (30.7) |
|  | Outside | 140 (33.6) |

\* Other include tools, medicine, pots and pans, household items

**Table 2.** Types of antibiotics and indications for use among traders in CBD

|  | Drug |  |  |  |  |
| --- | --- | --- | --- | --- | --- |
|  | Penicillins | Flagyl | Antifungals | Others | TOTAL |
| URTIs N (%) | 44 (33.1) | 18 (20.7) | 6 (46.1) | 26 (56.5) | 94 (33.7) |
| GIT N (%) | 27 (20.3) | 51 (58.6) | 3 (23.1) | 5 (10.9) | 86 (30.8) |
| Skin N (%) | 30 (22.6) | 8 (9.2) | 1 (7.7) | 5 (10.9) | 44 (15.8) |
| Fever/Pain N (%) | 11 (8.3) | 5 (5.7) | 0 (0.0) | 2 (4.3) | 18 (6.5) |
| Genital Discharge N (%) | 13 (9.7) | 4 (4.6) | 3 (23.1) | 7 (15.2) | 27 (9.7) |
| Other N (%) | 8 (6.0) | 1 (1.2) | 0 (0.0) | 1 (2.2) | 10 (3.5) |
| Number of respondents N (%) | 133 (47.67) | 87 (31.18) | 13 (4.65) | 46 (16.49) | 279 (66.91) |

Penicillins such as the tablets formulations of amoxicillin, penicillin V, flucloxacillin, and amoxicillin/clavulanic acid combination were the most (N= 133 (47.67%)) used in antibiotic self-medication. The next most self-medicated antibiotic was metronidazole (N=87 (31.18%)).

### Source of information and adherence to antibiotic dosage among traders in CBD

More than half of traders who practiced self-medication with antibiotics (N= 158 (55.2%)) had previously consulted pharmacists, while 17.8% (N=51) had consulted their doctors. Moreover, most of them (67.5% (193)) knew the quantity to buy from the pharmacist, and about a fifth (20.3% (N= 58)) indicated the use of a previous prescription.

About 80% (N=227) reported never changing the dosage, while 16.0% (N=45) said they sometimes changed the dosages. For those who admitted to changing dosage, the given reasons included feeling better (43.7% (N=28), unpleasant side effects (23.4% (N=15)), and worsening of conditions (17.2% (N=11)).

Most participants (44%, N= 125) indicated they stopped taking the antibiotics after the symptoms disappeared, while only 1.8% (N=5) stopped upon consultation with a doctor or pharmacist.

### Factors associated with self-medication with antibiotics (Table 4)

At the univariable level, females were 1.6 (95% CI; 1.03-2.40) times more likely than males to practice self-medication with antibiotics (p = 0.03), while those with a primary level of education were more likely (O.R. = 2.06, 95% CI; 0.83-5.11) than persons with no education to practice self-medication with antibiotics. Traders who had a tertiary level of education (OR= 0.37; 95% CI; 0.14-0.91) self-medicate with antibiotics compared to those with no education (p=0.01). Married couples were less likely (OR= 0.18, 95% CI; 0.54-1.3) than singles to practice self-medication. Persons who sold food items (OR= 4.32, 95% CI; 2.03-9.19), cosmetics (OR=2.75, 95% CI; 1.42-5.31), clothing (OR= 3.53, 95% CI; 1.43-5.31), and stationery (OR=1.70; 95% CI; 1.94-6.43) were more likely to self-medicate with antibiotics compared to those who sold other items (p <0.01).

**Table 3:**
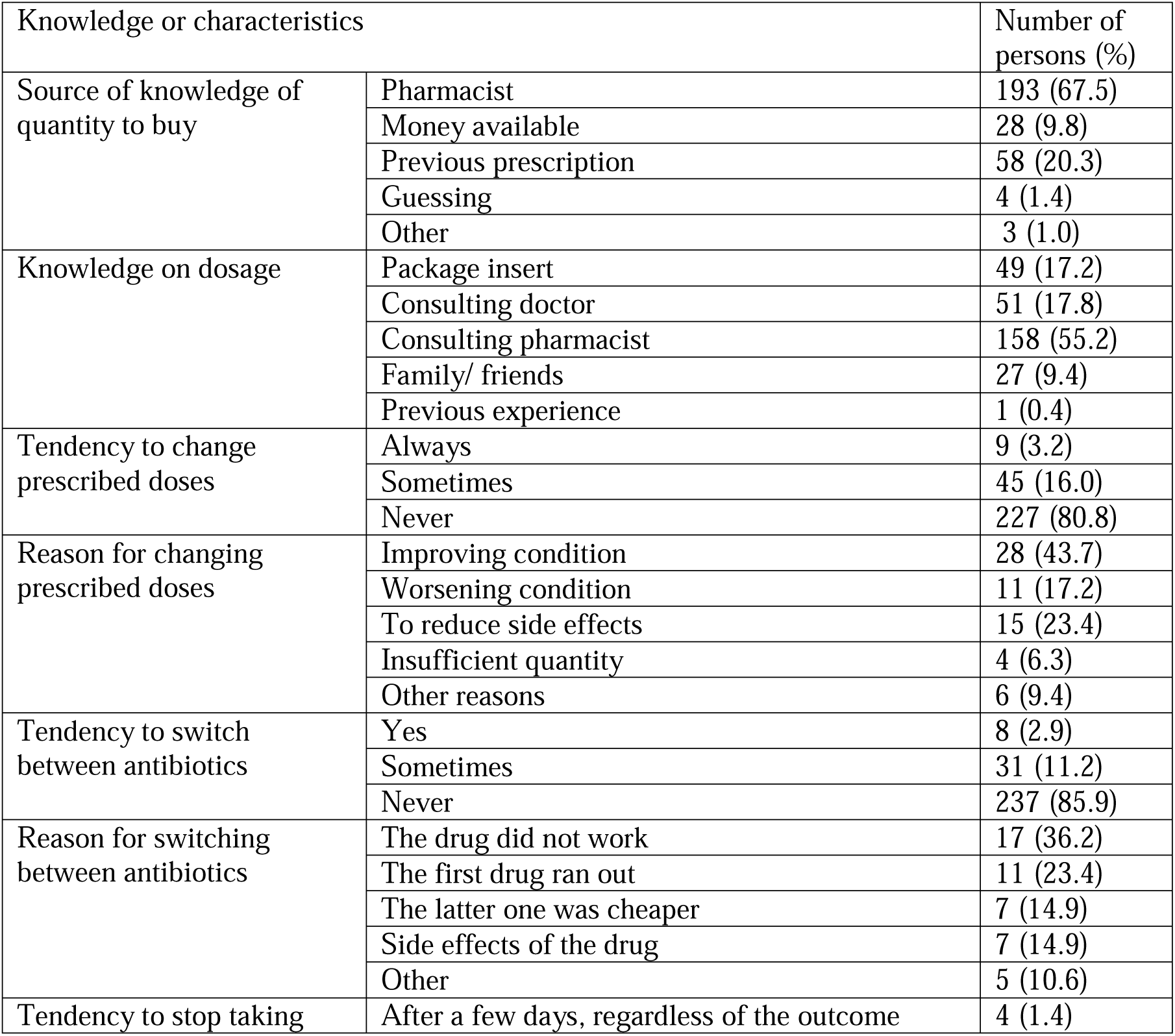
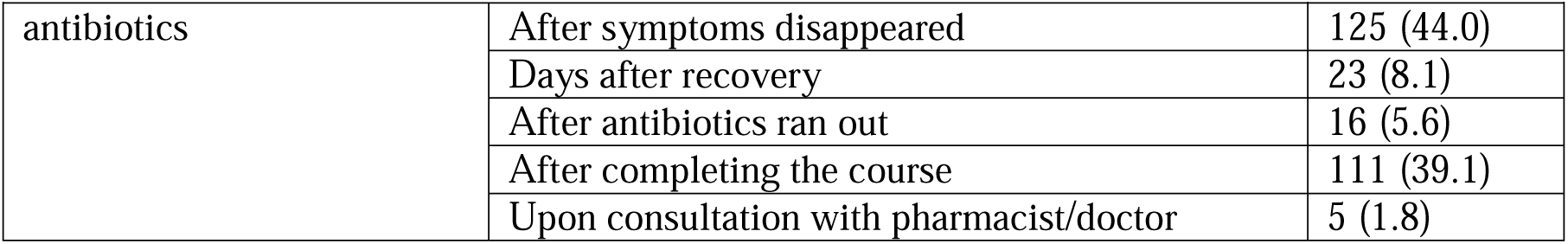
Knowledge and characteristics of antibiotic use among traders in the Central Business District of Accra, Ghana (N=417).

**Table 4:** Demographic Predictors of self-medication with antibiotics among traders in CBD ^a^ Excluded from the multivariate model because of nonsignificant effect at the univariate model. Source: Author’s Field Survey, December 2016

| Indicator |  | Antibiotic use |  | Unadjusted model |  | Adjusted model |  |
| --- | --- | --- | --- | --- | --- | --- | --- |
|  |  | Yes (%) | No (%) | OR (95%) | P value | OR (95%) | P value |
| Age (years)<sup>a</sup> | 31-45 | 123 (66.5) | 62 (33.5) | 1.05 (0.67-1.67) | 0.72 | NA |  |
|  | >45 years | 45 (70.3) | 19(29.7) | 1.26 (0.67-2.37) |  |  |  |
| Sex | Female | 171 (70.7) | 71(29.3) | 1.6 (1.03-2.40) | 0.03 | 1.36 (0.83-2.24) | 0.22 |
| Education | Primary | 76 (84.4) | 14 (15.6) | 2.06 (0.83-5.11) | 0.01 | 1.97 (0.78-4.97) | 0.15 |
|  | Secondary/Middle | 135 (62.5) | 81 (37.5) | 0.63 (0.23-1.34) |  | 0.63 (0.29-1.34) | 0.25 |
|  | Tertiary | 26 (49.1) | 27 (49.1) | 0.37 (0.14-0.91) |  | 0.39 (0.16-0.98) | 0.04 |
| Marital status<sup>a</sup> | Married/Cohabiting | 120 (62.8) | 71 (37.2) | 0.83 (0.54-1.3) | 0.18 | NA |  |
|  | Separated/Divorced | 13 (86.7) | 2 (13.3) | 3.19 (0.69-14.8) |  |  |  |
|  | Widowed | 17 (85.0) | 3(15.0) | 2.78 (0.77-10.01) |  |  |  |
| Place of trade<sup>a</sup> | Rented shop | 80 (67.2) | 39 (32.8) | 0.93 (0.56-1.57) | 0.75 | NA |  |
|  | Outside | 85 (63.9) | 48 (36.1) | 0.80 (0.49-1.33) |  |  |  |
| Items traded | Food | 54 (78.3) | 15 (21.7) | 4.32 (2.03-9.19) | 0.01 | 3.56 (1.70-7.45) | 0.01 |
|  | Cosmetics | 55 (69.6) | 24 (30.4) | 2.75 (1.42-5.31) |  | 2.61 (1.34-5.09) | 0.01 |
|  | Clothing | 100 (74.6) | 34 (25.4) | 3.53 (1.43-5.31) |  | 3.39 (1.88-6.12) | 0.01 |
|  | Stationary | 17 (58.6) | 12 (41.4) | 1.70 (1.94-6.43) |  | 2.01 (0.79-5.09) | 0.14 |

In the multivariate analysis, persons with tertiary education were less likely (aOR = 0.39, 95%CI; 0.16-0.98) to use antibiotics for self-medication compared to those with no education (p= 0.04). Persons who sold food items (aOR = 3.56, 95% CI; 1.70-7.45), cosmetics (aOR = 2.61, 95% CI; 1.34-5.09), and clothing (aOR= 3.39, 95% CI; 1.88-6.12) were more likely to use antibiotics for self-medication compared to those who sold other items.

## Discussion

The inappropriate use of oral antibiotics in lower and middle-income countries remains a challenge to achieving the third sustainable development goal, subsequently contributing to the increase in antibiotic resistance. Managing all forms of infectious diseases at health facilities largely depends on effective case detection and diagnosis through systematic screening and, ultimately, treatment with appropriate antibiotics. However, vulnerable populations such as traders tend to purchase antibiotics mostly from retail vendors to address self-diagnosed conditions without prescriptions. The study has found that the prevalence of antibiotic self-medication among traders in the CBD, Accra was 66.7%. The most commonly self-medicated are the Penicillins, with the treatment of upper respiratory infections being the main reason for self-medication. Whereas traders who sold food items, cosmetics, and clothes were more likely to self-medicate than other traders, those with higher education were less likely to self-medicate than those without any education.

A previous study that explored antibiotic self-medication among university students in Accra, Ghana, found the prevalence to be 21% [17]. The higher prevalence of 66.7% found in the present study among traders is consistent with the further finding of the effect of education on the tendency to self-medicate [18]. The high prevalence can be explained by the country being disproportionately affected by infectious diseases encouraging the use of antibiotics for self-medication, limited access to healthcare, and lack of antibiotic regulations [19, 20].

In our study, about 10% of traders had at least a tertiary level of education. This may lead to an assumption that the higher the educational level, the less likely it was to self-medicate with antibiotics. A study conducted in Serbia buttressed this assumption, reporting a self-medication prevalence of 46% among university students [21]. An attributable reason for the observed high prevalence in this study could be that although antibiotics are prescription-only medicines, regulatory measures are not strict enough to enforce them, making them easily accessible to such drugs [22]. In addition to these regulatory aspects, other differences in health care systems such as drug prices and reimbursement policies may also influence the public’s attitudes towards antibiotic use and self-medication [23].

This study revealed five main conditions for which antibiotics were used. Upper respiratory tract infections (URTIs), reported as cough, common cold, and sore throat, were the most common ones like those reported in studies in Nigeria, Kuwait, and Ethiopia [11, 15, 24]. A study on antibiotic use in children under five years in Kintampo Municipal Hospital, Ghana, also mentioned upper respiratory tract infections as the major indication for antibiotic use [25]. In this study, gastrointestinal disorders recorded as diarrhea and stomachache followed URTIs. This is consistent with findings in other studies [26, 27].

The problem with using antibiotics for diarrhea is that viruses cause most diarrhea cases, and hence it is not imperative to use antibiotics for such cases. This creates a condition for the development of resistance to antibiotics [27]. Similar to our findings, other studies also mentioned skin diseases and penile/urethral discharge as indications for antibiotics use [11].

Among the several available antibiotics, our findings identified two major antibiotics used for self-medication: penicillins and metronidazole. The choice of antibiotics from the penicillins group (especially flucloxacillin and amoxicillin) by most respondents in our study is in line with findings from other studies [27, 28]. The common reasons for the patronage of penicillins could be because they are cheap, easily accessible, have a good safety profile, and possesses broad-spectrum antimicrobial activity [22]. The reasons can further be explored using qualitative research. Also, even though antibiotics are supposed to be prescription-only medicines, they are easily accessible partly because regulatory measures are not strictly enforced. The frequency of metronidazole use as recorded in this study was high, compared to another study in Northern Nigeria which reported a value of 17.6% [27]. The difference could be due to socio-demographic variation between study participants. For research purposes, antifungals were included in the definition of antibiotics, and similar to our findings, it was found out in another study that antifungals were used chiefly for skin-related infections [29].

Education was an independent predictor for antibiotic self-medication among the traders. The odds of self-medication were lower among respondents with a higher level of education relative to traders with lower education. This parallels results from other studies where varying levels of education impacted the likelihood to self-medicate with antibiotics [6, 30]. This phenomenon may be attributed to the fact that educated persons are likely to have knowledge of the implications of antibiotic misuse and hence desist from the act. However, other studies dispute this fact, reporting that more educated people had a higher tendency to use antibiotics [31]. Contrary to findings in other studies [6, 32], this study did not find gender to be independently associated with self-medication with antibiotics.

Three significant sources of information on antibiotics for self-medication were identified: the pharmacist, the doctor, and package inserts. Pharmacists have also been identified in most studies as the primary source of information on drugs used in self-medication [33]. In their professional capacity and direct contact with patients, the community pharmacist is competent to provide sound advice on the medicines they supply and screen for conditions at the level of a primary healthcare provider [11, 34]. Hence, the role of community pharmacists in self–medication with antibiotics needs to be encouraged to promote responsible use of drugs. Consulting doctors, in other studies, was, however, the primary source of information compared to pharmacists [30]. The difference in these studies could be due to socio-demographic differences and health-seeking behaviors. A finding in this study was that the third common source of information was reading the package leaflet. Some studies mentioned reading pamphlets as a source of information, but the participants were university students and lecturers [29, 35]. Friends and family members were also revealed to be a source of information in this study, and this has been reported in studies in Egypt [7] and among adolescents in Kuwait [24]. This may reflect the belief in the value of other people’s experiences and the views of older adults.

## Limitations

Antibiotic self-medication was self-reported, and some participants may have not voluntarily reported the practice because they knew it was frowned upon, and there is public education against self-medication. Therefore, it is conceivable that the prevalence determined in this study was an underestimation and the true prevalence is much higher.

The lack of a complete list of traders and their shops made it impossible to target a representative sample of the traders due to the majority of participants being mobile. This limits the generalizability of the study’s findings; however, data were collected mainly in three of the busiest market centers in Accra. Finally, there may be recall bias since respondents were asked to recount their antibiotic use in the past year.

## Conclusion

The prevalence of self-medication with antibiotics was high among the traders, and penicillins are the most common antibiotics used in self-medication. Higher education and the type of items sold were independent predictors of self-medication practice among the respondents.

We recommend that a holistic approach should be used in addressing self-medication with antibiotics and this would involve strengthening drug regulatory measures, organizing health education programs on the dangers of self-medicating with antibiotics and the implications of antibiotic resistance, and empowering community pharmacists operating in the CBD to provide one-on-one education for traders who call to procure analgesics and antibiotics.

## Data Availability

All data produced in the present study are available upon reasonable request to the authors

